# Use of Paired Vagus Nerve Stimulation in Mobilization of Patients with Prolonged Disorders of Consciousness: Protocol for a Single Cohort Open Label Interventional Study

**DOI:** 10.1101/2025.07.24.25332147

**Authors:** Jenna Tosto-Mancuso, Jessica Polizzi, Neha S. Dangayach, Aidan Rogers, Christopher P. Kellner, Madeline Fields, Wilber Parada Iraheta, Sofia Barchuk, Raphael Kesselman, David Putrino

**Affiliations:** Department of Rehabilitation and Human Performance, Icahn School of Medicine at Mount Sinai; Department of Neurosurgery, Icahn School of Medicine at Mount Sinai; Department of Neurology, Icahn School of Medicine at Mount Sinai

**Keywords:** Neural Stimulation, Disorders of Consciousness, Neurorehabilitation, Brain Injury, Chronic, Nerve Stimulation, Vagus

## Abstract

**Introduction:** Severe acquired brain injury (SABI) is a significant global health concern that can lead to prolonged disorders of consciousness (pDOC). Rehabilitation in the current standard of care is limited to sensory stimulation interventions exclusive to the acute and subacute phases of injury. This work seeks to demonstrate the safety, feasibility, and preliminary efficacy use of the novel pairing of neural stimulation (taVNS) and robotic tilt table mobilization (RTTM) in the rehabilitation of patients with pDOC.

**Methods and Analysis:** Fifteen (15) participants with pDOC (>3 months since onset of disorder of consciousness) will participate in three rehabilitation intervention phases: 1) observation of standard of care; 2) intervention phase (2a: RTTM only and 2b: RTTM plus taVNS);and 3) long term follow up at 3, 6, and 12 months post interventions. Primary outcomes will include reporting of adverse and serious adverse events and adverse and serious adverse reactions; they will be collected throughout the study duration. Secondary feasibility outcomes will include number of sessions completed, time on therapy task completed, number of steps taken with robotic stepping, maximal limb loading or the maximal amount of weight distribution through single limb stance of the robotic gait cycle, and maximal tilt angle and will be collected each study session. Exploratory outcomes will include the Coma Recovery Scale-Revised (CRS-R), Glasgow Coma Scale (GCS), and Glasgow Outcomes Scale- Extended (GOSE), Nociceptive Coma Scale-Revised (NCS-R) and hemodynamics (blood pressure [BP], heart rate [HR], oxygen saturation [SpO2]) assessments to be collected at baseline, immediately following phase 1; immediately following phase 2a, immediately following phase 2b and at long term follow up time points. Additionally, quantitative electroencephalography (qEEG) will be collected at the end of phase 1, end of phase 2 (end of study intervention) and at the each longitudinal follow up study visits. The aim will be to evaluate proof of concept in this safety feasibility trial.

**Ethics and Dissemination:** The study protocol has been approved the Program for the Protection of Human Subjects (PPHS) at the Icahn School of Medicine at Mount Sinai (Study 24-01339)

**Registration:** NCT06930716

**Article Summary:** *Strengths and limitations of this study:*

- This work seeks to evaluate a novel paired neuromodulation and physical therapy intervention for patients with prolonged Disorders of Consciousness (pDOC)
- This work exclusively will evaluate the benefits of rehabilitation and subsequent outcomes in a disparate population with limited treatment options
- This trial is not randomized, limiting rigor of the study design
- This trial is unpowered, limiting generalizability to the larger pDOC population
- Future work will seek to address these limitations

## Introduction and Rationale

Prolonged disorders of consciousness (pDOC) are defined as disorders of consciousness (DOC) lasting >1 year post injury for patients with traumatic brain injury (TBI) and >3 months post injury for patients with non-TBI.^1^ At present there are limited interventions that reliably lead to enhanced prognosis.^1^ The rehabilitation process throughout the continuum of care for patients with pDOC necessitates restorative strategies to facilitate arousal and functional recovery along with coordinated medical management.^2^ Rehabilitation interventions for patients with DOC and pDOC have evolved in the past decade, with an emerging body of evidence highlighting the benefits of rehabilitation intervention even in the acute stages of coma and DOC.^1–3^ Use of coma stimulation protocols including systematic introduction of somatosensory stimuli, progressive mobilization, neural stimulation and neuromodulation have been introduced as a standard of care for acute and subacute physical therapy respectively. ^4–6^ Implementation of progressive upright verticalization and mobilization has been shown to improve both short- and long-term outcomes, arousal, orthostatic tolerance and hemodynamic stability. ^7–9^ Robotic tilt table mobilization (RTTM) (ErigoPro, Hocoma) has been shown to be safe, feasible, and well tolerated in patients with DOC. ^10–12^ Evaluation of this form of mobilization, RTTM with passive verticalization, robotic enabled lower extremity stepping has been shown to improve short- and long-term functional and cognitive outcomes in patients with DOC when compared to conservative therapy alone.^8^

In addition to physical interventions like RTTM, the use of invasive and non-invasive neural stimulation has been investigated as an adjunct to rehabilitation for patients with DOC. ^1,2^ Among these, vagus nerve stimulation (VNS) has demonstrated the potential to maximize recovery across acquired brain injuries in both acute and chronic phases.^13–15^ Pairing VNS with quality rehabilitation has demonstrated benefits in other neurologic populations including motor recovery in chronic stroke. ^16^ Through modulation of acetylcholine and norepinephrine transauricular vagus nerve stimulation (taVNS), a method of non-invasively stimulating the vagus nerve by applying electrical stimulation at the ear, has been identified as safe and potentially effective in patients with acute and chronic DOC promoting neurobehavioral changes and increased functional connectivity on imaging. ^14,17–19^ Such findings have been seen in functional magnetic resonance imaging (fMRI) in key neural regions responsible for arousal and consciousness including the thalamus, ventral medial prefrontal cortex, superior temporal gyrus, and posterior cingulate. ^13,14^ While there is data to support the individual utility of these modalities, no work to date has investigated the benefits of pairing taVNS and RTTM to maximize functional recovery in patients with pDOC. This work will report on the safety, feasibility, and preliminary short- and long-term outcomes of RTTM with simultaneously paired taVNS for subacute and chronic acquired brain injury patients with pDOC.

## Methods and analysis

### Participants

This single center cohort pilot feasibility study will seek to rapidly evaluate the safety, feasibility, and preliminary efficacy of paired transauricular vagus nerve stimulation (taVNS) and robotic tilt table mobilization (RTTM) in the physical therapy rehabilitation of patients with pDOC. This study protocol was developed in accordance with SPIRIT reporting guidelines.^20^ The study will be conducted at the Abilities Research Center at the Icahn School of Medicine at Mount Sinai Health in New York City, United States. Fifteen (15) participants will be recruited from the Mount Sinai Health System (MSHS) from various programs that span established, interdisciplinary collaborations between the Departments of Rehabilitation and Human Performance, Neurosurgery and Neurology at the MSHS as well as the broader community. As this is a safety feasibility study, study subjects will not be randomized and study team members will not be blinded. While no power analysis was completed as this is a pilot feasibility trial, the number of participants to be enrolled in this trial are reflective of trials of similar study design.

The study team will screen patients receiving clinical care at the MSHS for participation in this study. Those who contact the study team with an interest in participation will be screened by the study physicians and study team members. The study will enroll patients who meet the following inclusion criteria: patients with a diagnosis of pDOC (includes coma, vegetative state, unresponsive wakefulness syndrome, minimaly conscious state, minimally conscious state+, and minimally conscious state-) as defined as a DOC greater than 3 months post onset and patients deemed medically safe to participate in physical therapy (PT) as evaluated by one of the physicians. Exclusion criteria will preclude patients who have emerged from MCS (Coma Recovery Scale- Revised (CRS-R) score 6 on Motor Function scale and/or 2 on Communication Scale). Patients medically unsafe for participation in PT as evaluated by one of the study physicians (including but not limited to those receiving intravenous sedation, those with integumentary breakdown or known pressure injuries, those with cardiovascular or cerebrovascular conditions precluding initiation of physical therapy (i.e. uncontrolled intracranial pressure, severe symptomatic orthostatic hypotension, etc.) Patients with DOC less than 3 months post onset will also be excluded. Additionally, patients who do not meet technical requirements of the RTTM device will be excluded (patients weighing greater than 135 kg, length of legs below 75 cm or above 100 cm, and fixed contractures of lower extremity including hip, knee, ankle, or foot). Given the medical complexities of patients with pDOC, the study team will require patients to have medical clearance from one of the study physicians to participate. Should the study physicians or study team feel the patient is no longer medically appropriate for participation in the trial or if the study physician and/or study team feel the patient is unduly distressed, the study protocol will be stopped, and the patient will be withdrawn from the trial. Similarly, if the patient requests to stop study participation, they will be withdrawn from the trial.

To optimize retention, participants will be scheduled at their convenience during normal outpatient office hours. The study team will communicate with the patient and care partner regarding the overall study plan and design. The study physical therapists will work closely with the study physicians to ensure the patient remains medically appropriate for participation in the study intervention. The study therapists and study assistant will coordinate with the patient and caregivers to ensure support is provided for scheduling transportation to and from study visits. To enhance retention, we will implement regular follow-up calls and maintain open communication with participants and their families, providing support and addressing concerns throughout the study duration. The main barriers to participation include time commitment of the intervention and travel to the study sites. To minimize these barriers, we will work to coordinate and conduct study activities at times that are convenient for the study participants. We will also provide renumeration to subjects to encourage adherence and retention.

### Study Interventions

Once study team members have completed written informed consent with a participant’s legally authorized representatives (LAR), the participant will complete three study phases: Timepoint(T) 1- a baseline observation of standard of care, T2- intervention, and T3 longitudinal follow up. While partaking in the study, participants will be encouraged to continue with their established standard of care including any medications, outside therapy, or other medical treatments.

At baseline T1, participants will be assessed using Coma Recovery Scale-Revised (CRS-R), Glasgow Coma Scale (GCS), and Glasgow Outcomes Scale- Extended (GOSE). In T1, participants will continue with 4 weeks of their current standard of care. During T1, no study interventions will be implemented. At completion of T1, the CRS-R, GCS, and GOSE will be reassessed.

T2 will be divided into two subphases to account for ramp up intervention consistent with clinical practice. T2a (RTTM only) will entail a 4-week mobilization only phase to ramp up patient tolerance to mobilization. Participants will complete 8 sessions of physical therapy (PT) over 4 weeks, receiving progressive verticalization and mobilization using a robotic tilt table with robotic stepping (Erigo; Hocoma, Switzerland). Pain will be assessed with the Nociceptive Coma Scale-Revised (NCS-R) and hemodynamics (blood pressure [BP], heart rate [HR], oxygen saturation [SpO2]) at the start of treatment sessions prior to mobilization, at stepping initiation, at tilt initiation, at 5-minute intervals while at maximal tilt, and post session. Stepping parameters will be set for a target of 30-60 steps per minute. The initial verticalization angle will be increased from 0° to 35-45° to assess orthostatic tolerance. Tilt angle will then be increased to a maximal tilt of 60-90° within 10 minutes of stepping initiation. Maximal tilt angle will be maintained for up to 20 minutes. At the end of each session the participant will be progressively returned to 0° over a period of 10 minutes, lowered in 10° increments.

Primary outcomes including Adverse events (AE), serious adverse events (SAE), the NC and reactions will be reported. An AE will be defined as any negative or unintended non-serious event occurring during the trial. A SAE will be defined as any negative or unintended event that results in a life-threatening injury, hospitalization, or resultant persistent or significant disability or death - whether related to the study intervention or not. An adverse reaction (AR) will be defined as above, directly related to the study intervention. Severe adverse reactions (SAR) will be defined as SAE but directly related to the study intervention (taVNS+RTTM). Feasibility outcomes including training time on task or the amount of actual intervention time received will be reported. Training data derived from the Erigo software will be included in data collection including number of steps taken with robotic stepping, maximal limb loading or the maximal amount of weight distribution through single limb stance of the robotic gait cycle, and maximal tilt angle will be collected each session. At completion of T2a, the CRS-R, GCS, and GOSE will be reassessed as a midpoint of the intervention phase.

T2b will include completion of 8 sessions of PT over 4 weeks, receiving the paired taVNS + RTTM intervention. The mobilization protocol will continue as described, with the addition of taVNS (VaguStim, Tel Aviv) administered at the ear immediately prior to initiation of tilt. Stimulation parameters (square waveform; pulse width: 200- 250 us; frequency: 20 Hz; amplitude: maximum of 1.5 mA) will be used, consistent with previously reported parameters for the use of taVNS in the cohort. ^14,17,18^ The study protocol will be stopped if at any point the participant demonstrates a heart rate of less than 40 or greater than 150 bpm, orthostatic hypotension (decrease in blood pressure of less than 20mmHg systolic or less than or equal to 10mmHg diastolic from baseline), oxygen saturation less than 90%, traumatic dislodgement of a device (i.e. tracheal cannula, intravenous or arterial line, or urinary catheter) or demonstrates a significant change in NCS-R score. ^21^ Upon completion of T2b, participants will be reassessed with CRS-R, GCS, and GOSE.

Subsequent longitudinal follow up (T3) will include three additional study visits for assessment at 3-, 6-, and 12-months post intervention using the CRS-R, GOSE, and GCS. Across all study phases, secondary healthcare utilization, including re-admission rates, number of physician follow up visits and emergency room visits, will be captured from the patient electronic medical record (EMR) and reported.

### Quantitative Electroencephalography

In addition to behavioral and functional outcomes conducted throughout the study as previously detailed, participants will complete the CRS-R while simultaneously being evaluated with quantitative electroencephalography (qEEG) to evaluate cortical response to the intervention.^22,23^ The qEEG assessments will be completed at the end of T1 (end of standard of care observation/beginning of intervention), end of T2 (end of study intervention) and at the each longitudinal follow up study visit (3, 6, 12 months post-intervention). Steady state assessment as well as event related potentials (ERPs) will be collected using the Enobio 32 (Neuroelectrics, Barcelona). The Enobio 32 is a wireless thirty-two electrode channel EEG sensory system that allows for the capture of neuroelectrical activity from the brain. Location of electrodes of the Enobio 32 correspond to the 32-channel montage of the international 10-20 system. Wet electrodes will be used for data acquisition and affixed to participants’ heads via a neoprene headcap in said international 10-20 system formation. All electrode signals will be referenced to a linked pair of electrodes located on the earlobe. The data will be sampled at a rate of 500Hz (with optional downsampling during postprocessing) and with electrode impedance <1 GΩ. Bandpass filters will be applied post processing at 0.01-100Hz consistent with established literature. Approximately 20-25 minutes will be required for donning and setup of EEG. EEG quality will be assessed based on visual appearance, consistency of waveforms, line noise (uV^2^) (60hz+/-1hz) and main noise (uV^2^)(1-40hz) and mean waveform offset. Further, signal quality metrics derived from the software of the Enobio32 system will be used to guide signal quality evaluation. A ten-minute steady state sample will be collected followed by presentation of the Coma Recovery Scale-Revised.

### Analysis

A robust data collection and analysis plan will be deployed to ensure best practices. Primary outcomes include adverse event reporting and will be captured subjectively and recorded as number of events. Details of the events will also be captured. Primary outcomes will be represented with descriptive statistics including mean number of instances. Secondary outcomes including number of sessions completed, time on task, number of steps taken with robotic stepping, maximal limb loading or the maximal amount of weight distribution through single limb stance of the robotic gait cycle, and maximal tilt angle will be represented with descriptive statistics (mean [range]) to describe central tendency within each phase of the intervention. Paired t-tests will be used to compare baseline and post intervention CRS-R, GCS, and GOSE scores respectively within each phase. Tests of multiple comparisons will be used to evaluate significant differences between groups longitudinally. Additionally, descriptive statistics and repeated measures ANOVA will be used to compare CRS-R, GCS, GOSE scores and interventional data (number of steps, orthostatic hemodynamics, mean tilt, max limb loading, max tilt angle and the Nociceptive Coma Scale-Revised (NCS-R).

qEEG data analysis will include event related potential (ERP) amplitude and latency analysis and power spectral analysis, aswell as, coherence analysis of steady state data, and brain symmetry indices. Post processing of the data samples will include the use of a bandpass filter (bandpass; 0.1-100 Hz) as well as data cleaning to ensure artifact removal and noise reduction. Data will subsequently be re-referenced to the common average reference. Notch filtering will be employed to reduce line noise. An independent component analysis (ICA) will be used to remove artifacts, which will vary by subject. Data will be aligned with events including the 5 second ramp period and marked CRS-R test item introduction and task completion. Subsequently, data will be separated into epochs based on cadence between stimulus introductions (5 second ramp, 60 second stimulus duration, 5 second ramp down). Mean power will be calculated over each epoch over a frequency range of 0.5-50Hz using windowed Fast Fourier Transform (FFT) spectral analysis. Calculation of the mean power at each frequency band will be calculated across all scalp electrodes, creating an average scalp spectra. From the average scalp spectra, absolute power will be summed across the four primary frequency bands: delta (1-4Hz), theta (4.1-8Hz), alpha (8.1-12.5Hz), and beta (12.6-30Hz). Following transformation of the EEG signal to frequency domains using FFT as described above, power spectral density analysis (PSD) will be used to further represent the signal as a function of frequency. PSD will be calculated as amplitude squared of the frequency band derived from the Fourier transform.^10 22,24^Power spectral density plots with electrode mapping will be used to visualize distribution of neuronal activity across frequency bands. Topographic maps will be used to visualize the distribution of frequency bands. Brain Symmetry Index (BSI) will be calculated as a measure of interhemispheric power asymmetry, evaluating differences in power spectra in the left and right hemispheres, respectively. BSI is commonly used to evaluate asymmetry and in the context of qEEG, allows for quantification and subsequent visualization of temporal and spatial data.^10,25,26^ BSI will be calculated from steady state data and ERP’s from each stimulus response during the CRS-R. BSI will be calculated using methods previously reported by van Putten to allow for increased sensitivity to interhemispheric asymmetry.^12^ Functional connectivity will be analyzed using coherence analysis to evaluate the synchronization and phase relationship of cortical activity among distinct regions.^23^ Coherence analysis will seek to evaluate the phase and amplitude variance among electrodes to better understand synchronization. ^27^

## Ethics and dissemination

The study team will require a patient to have an identified care partner or legally authorized representative (LAR) to consent to participation and to provide support to and from study visits. The study team will maintain the heterogeneity of the sample and ensure a real-world sample of the diverse patients served at this large urban health care system by recruiting from existing clinical infrastructure for this study. Informed consent will be obtained from all participants’ LARs in accordance with approval received from the Program for Protection of Human Subjects (PPHS) at the Icahn School of Medicine at Mount Sinai (STUDY 24-01339). Any requests for changes to this protocol will be approved by PPHS Institutional Review Board (IRB) prior to implementation.

Study data will be collected and managed using REDCap electronic data capture tools hosted at the Icahn School of Medicine at Mount Sinai. ^28,29^ REDCap (Research Electronic Data Capture) is a secure, web-based software platform designed to support data capture for research studies, providing 1) an intuitive interface for validated data capture; 2) audit trails for tracking data manipulation and export procedures; 3) automated export procedures for seamless data downloads to common statistical packages; and 4) procedures for data integration and interoperability with external sources.

Dissemination of the proposed work will be planned for publication upon completion of data analysis and final manuscript development.

## Data Availability

No data has been collected as this is a protocol.

## Author Contributions

JTM and DP conceptualized the trial; JTM, JP, WPI, ND, CK, and MF contributed to rehabilitation clinical protocol development and will carry out study interventions; ND, CK, MF, and SB contributed to the medical protocol development; JTM, AR, and MF contributed to the data analysis plan; JTM and DP designed the statistical analysis plan; All authors contributed to manuscript development and provided final approval.

## Funding statement

This work was supported by The Foundation for Physical Therapy Research Magistro Family Foundation Grant Number 1333107.

Foundation for Physical Therapy Research

3030 Potomac Ave., Suite 110

Alexandria, VA 22305-3085

1-800-875-1378

## Trial Registration Number

NCT06930716

## Acknowledgements

The study team wishes to acknowledge our Department Chairs Dr. Joseph Herrera and Dr. Joshua Bederson for their support of this work. Additionally, we wish to thank the participants, their care-partners, and families for participation in this trial.

